# Effectiveness of an Impella versus intra-aortic balloon pump in patients who received extracorporeal membrane oxygenation

**DOI:** 10.1101/2024.03.28.24305040

**Authors:** Yuji Nishimoto, Hiroyuki Ohbe, Jun Nakata, Toru Takiguchi, Mikio Nakajima, Yusuke Sasabuchi, Toshiaki Isogai, Hiroki Matsui, Yukihito Sato, Tetsuya Watanabe, Takahisa Yamada, Masatake Fukunami, Hideo Yasunaga

## Abstract

**Background:** Previous studies have suggested that left ventricular (LV) unloading with an intra-aortic balloon pump (IABP) or percutaneous ventricular assist device (Impella) in combination with extracorporeal membrane oxygenation (ECMO) is associated with lower mortality; however, it is unclear which is better. This study aimed to evaluate the effectiveness of LV unloading with an Impella versus IABP on in-hospital mortality and other clinical outcomes.

**Methods:** Using the Japanese Diagnosis Procedure Combination database from September 28, 2016, to March 31, 2022, we identified inpatients who received an Impella or IABP in combination with ECMO (ECPella or ECMO+IABP group, respectively). The primary outcome was in-hospital mortality and the secondary outcomes were the length of hospital stay, length of ECMO, total hospitalization cost, and complications. Propensity score matching was performed to compare the outcomes between the groups.

**Results:** Of 14,525 eligible patients, 603 (4.2%) received ECPella and 13,922 (96%) received ECMO+IABP. After propensity score matching, there was no significant difference in in-hospital mortality between the two groups (58.9% versus 56.6%; risk difference, 2.3%; 95% confidence interval, −3.9% to 8.5%). The ECPella group had a longer hospital stay, higher total hospitalization cost, and more frequent major bleeding, vascular complications, and renal replacement therapy during hospitalization than the ECMO+IABP group.

**Conclusions:** This nationwide inpatient database study showed that ECPella was not associated with a survival benefit but was associated with a longer hospital stay, higher total hospitalization cost, and more complications than ECMO+IABP.

## Introduction

Venoarterial extracorporeal membrane oxygenation (ECMO) provides strong hemodynamic support to patients with cardiogenic shock through retrograde aortic blood flow in addition to oxygenation;^1^ however, even the latest evidence does not support a survival benefit with the routine use of ECMO in patients with cardiogenic shock.^2,3^ Potential reasons may include bleeding complications, peripheral vascular ischemia, pulmonary edema, and myocardial ischemia induced by an increased left ventricular (LV) afterload due to a strong retrograde aortic blood flow.^1^

Previous studies have suggested that LV unloading with an intra-aortic balloon pump (IABP) or percutaneous ventricular assist device (Impella) in combination with ECMO is associated with a lower mortality.^4–8^ Those studies primarily focused on the effectiveness of LV unloading with an IABP or Impella, using patients who received ECMO alone as controls. The IABP reduces the LV afterload and provides indirect LV unloading through a negative systolic pressure in the descending aorta,^9^ whereas the Impella can directly decrease the LV overload and restore the pulmonary flow in patients with ECMO who present with severe LV dysfunction.^10^ No randomized controlled trial compared the outcomes of using the Impella versus IABP in combination with ECMO. A recent meta-analysis of seven small-scale observational studies showed no survival benefit of ECPella as compared to ECMO+IABP; however, those studies had a high heterogeneity and the results of the meta-analysis were underpowered to assess the comparative effectiveness.^11^ It remains unclear whether differences in the device characteristics between the IABP and Impella affect the clinical outcomes in patients with ECMO who require LV unloading. Therefore, the present study aimed to evaluate the effectiveness of LV unloading with an Impella versus IABP on the in-hospital mortality and other clinical outcomes, using a nationwide inpatient database in Japan.

## Methods

### Design and Ethical statement

This was a retrospective cohort study using an inpatient administrative database, and the study conformed to the RECORD reporting guidelines.^12^ This study was conducted in accordance with the amended Declaration of Helsinki and was approved by the Institutional Review Board of The University of Tokyo (approval number, 3501-(5); 19 May 2021). Because the data were anonymous, the Institutional Review Board waived the requirement for informed consent.

### Data source

We used the Japanese Diagnosis Procedure Combination inpatient database, which contained administrative claims data and discharge abstracts from more than 1,500 acute care hospitals and covers approximately 90% of all tertiary emergency hospitals in Japan.^13^ The database includes the following patient-level data for all hospitalizations: age, sex, diagnoses (main diagnosis, admission-precipitating diagnosis, most resource-consuming diagnosis, second-most resource-consuming diagnosis, comorbidities present at admission, and complications arising after admission) recorded with the *International Classification of Diseases, 10th Revision (ICD-10)* codes, daily procedures recorded using Japanese medical procedure codes, daily drug administration, and discharge status.^13^ A previous validation study showed that the specificity of the recorded diagnoses in the database exceeded 96%, the sensitivity of the diagnoses ranged from 50% to 80%, and the specificity and sensitivity of the procedures both exceeded 90%.^14^

### Study population

We identified all patients who received ECMO during hospitalization from September 28, 2016, to March 31, 2022. September 28, 2016, was the date when the Impella was approved for reimbursement under the national health insurance in Japan. We excluded patients aged <18 years and those who had received neither an Impella nor IABP within 2 days before or after the ECMO initiation. All patients were followed up until they died or were discharged from the hospital.

### Treatment groups

Patients who received an Impella within 2 days before or after the ECMO initiation were defined as the ECMO+Impella (ECPella) group. Patients who received IABP within 2 days before or after the ECMO initiation were defined as the ECMO+IABP group. Patients who received both an Impella and IABP within 2 days before or after the ECMO initiation were assigned to the group with the later initiation date of the Impella or IABP, or defined as the ECPella group if the Impella or IABP was initiated on the same day.

### Outcomes

The primary outcome was in-hospital mortality. The secondary outcomes were the length of hospital stay, length of ECMO, total hospitalization cost, and complications including major bleeding, an ischemic stroke, vascular complications, and renal replacement therapy during hospitalization. Major bleeding was defined as the presence of either intracranial bleeding, intraspinal bleeding, pericardial hematomas, intra-abdominal or retroperitoneal hematomas, intra-articular bleeding, intraocular bleeding, or compartment syndrome, which was in accordance with the International Society of Thrombosis and Haemostasis definition.^15^ Ischemic stroke was defined as the presence of cerebral infarction or transient ischemic attack. Vascular complications were defined as an injury to a blood vessel, noncoronary artery dissection, acquired arteriovenous fistula, acute limb thrombosis, and hemorrhage and/or hematoma following a circulatory system procedure.^16^ Those outcomes are defined by the ICD-10 codes listed in **Supplemental Table S1**.

### Covariates

The covariates were the fiscal year upon admission, age, sex, smoking history, body mass index upon admission, Japan Coma Scale upon admission,^17^ Charlson comorbidity index score, comorbidity of peripheral vascular diseases, physical function measured by the Barthel index score upon admission,^18^ cognitive function before admission, home medical care before admission, place before admission, ambulance use, primary diagnoses upon admission, extracorporeal cardiopulmonary resuscitation (ECPR), interventions (percutaneous coronary intervention, coronary artery bypass grafting, surgical valve procedures, or percutaneous valve procedures) before ECMO initiation, organ failure support on the day of the ECMO initiation other than ECMO and mechanical ventilation,^19^ and hospital characteristics (teaching hospital, tertiary emergency hospital, and annual hospital volume of ECMO). The primary diagnosis upon admission was defined when it appeared as a main diagnosis, admission-precipitating diagnosis, most resource-consuming diagnosis, or second most resource-consuming diagnosis^13^, and is defined by the ICD-10 codes listed in **Supplemental Table S1**. ECPR was defined as receiving chest compressions on the same day of the ECMO initiation.

### Statistical analysis

We performed a propensity score-matching analysis to compare the outcomes between the ECPella and ECMO+IABP groups.^20^ A multivariable logistic regression model using all the covariates listed in **Table 1** was employed to compute the propensity scores for the patients in the ECPella group. Then, we performed a one-to-one nearest-neighbor matching that randomly selected a patient from the treatment group and subsequently paired that patient with the patient in the control group with the closest propensity score.^21^ In the present study, each time a patient in the ECPella group was identified, the one-to-one nearest-neighbor matching without replacement was performed for the estimated propensity scores using a caliper width set at 20% of the standard deviation of the propensity scores in the patients in the ECMO+IABP group for the exact same calendar year and month.^20^ To assess the performance of the matching, the covariates were compared using standardized differences, with absolute standardized differences of ≤10% considered to denote negligible imbalances between the two groups.^22^ After the propensity score matching, the primary and secondary outcomes for the two groups were assessed through a generalized linear model accompanied by cluster-robust standard errors with hospitals as the clusters. Differences and their 95% confidence intervals were calculated with generalized linear models using the identity link function, irrespective of the outcome types. For the 60-day in-hospital mortality, we also generated Kaplan–Meier curves and performed log-rank tests in the matched cohort.

**Table 1.**
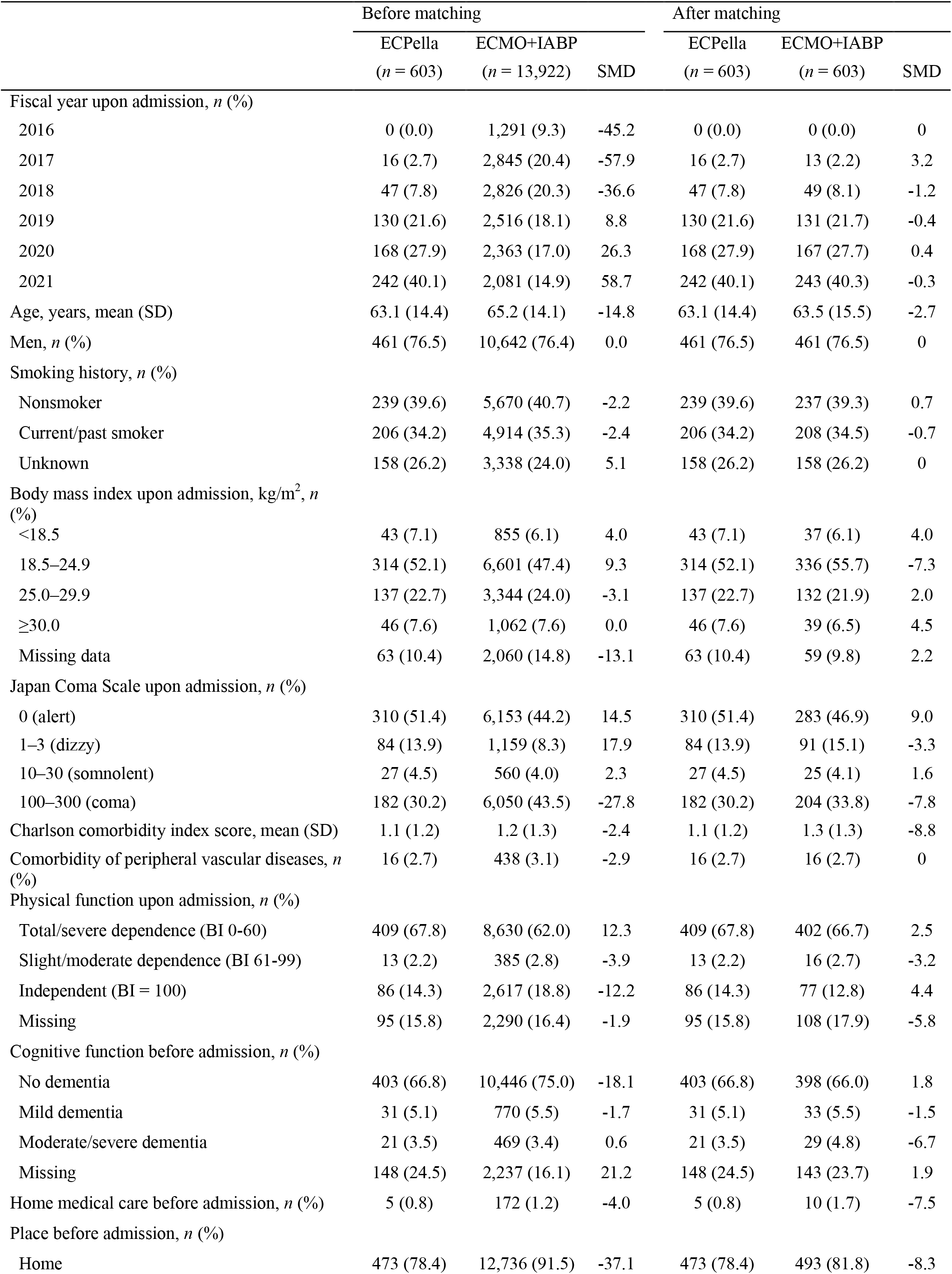
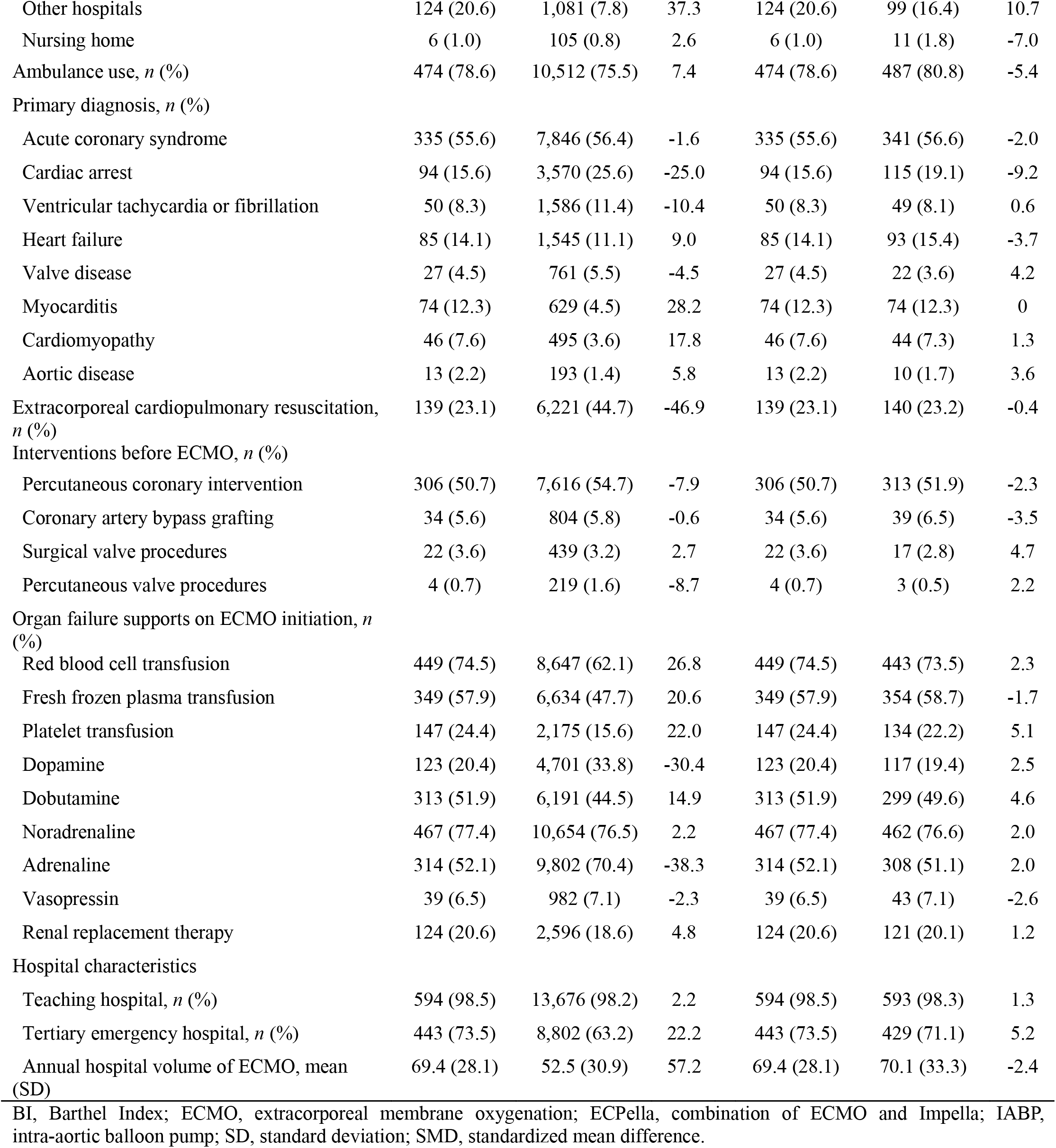
Patient characteristics.

|  | Before matching |  |  | After matching |  |  |
| --- | --- | --- | --- | --- | --- | --- |
|  | ECPella<br>(*n* = 603) | ECMO+IABP<br>(*n* = 13,922) | SMD | ECPella<br>(*n* = 603) | ECMO+IABP<br>(*n* = 603) | SMD |
| Fiscal year upon admission, *n* (%) |  |  |  |  |  |  |
| 2016 | 0 (0.0) | 1,291 (9.3) | -45.2 | 0 (0.0) | 0 (0.0) | 0 |
| 2017 | 16 (2.7) | 2,845 (20.4) | -57.9 | 16 (2.7) | 13 (2.2) | 3.2 |
| 2018 | 47 (7.8) | 2,826 (20.3) | -36.6 | 47 (7.8) | 49 (8.1) | -1.2 |
| 2019 | 130 (21.6) | 2,516 (18.1) | 8.8 | 130 (21.6) | 131 (21.7) | -0.4 |
| 2020 | 168 (27.9) | 2,363 (17.0) | 26.3 | 168 (27.9) | 167 (27.7) | 0.4 |
| 2021 | 242 (40.1) | 2,081 (14.9) | 58.7 | 242 (40.1) | 243 (40.3) | -0.3 |
| Age, years, mean (SD) | 63.1 (14.4) | 65.2 (14.1) | -14.8 | 63.1 (14.4) | 63.5 (15.5) | -2.7 |
| Men, *n* (%) | 461 (76.5) | 10,642 (76.4) | 0.0 | 461 (76.5) | 461 (76.5) | 0 |
| Smoking history, *n* (%) |  |  |  |  |  |  |
| Nonsmoker | 239 (39.6) | 5,670 (40.7) | -2.2 | 239 (39.6) | 237 (39.3) | 0.7 |
| Current/past smoker | 206 (34.2) | 4,914 (35.3) | -2.4 | 206 (34.2) | 208 (34.5) | -0.7 |
| Unknown | 158 (26.2) | 3,338 (24.0) | 5.1 | 158 (26.2) | 158 (26.2) | 0 |
| Body mass index upon admission, kg/m<sup>2</sup>, *n* (%) |  |  |  |  |  |  |
| <18.5 | 43 (7.1) | 855 (6.1) | 4.0 | 43 (7.1) | 37 (6.1) | 4.0 |
| 18.5–24.9 | 314 (52.1) | 6,601 (47.4) | 9.3 | 314 (52.1) | 336 (55.7) | -7.3 |
| 25.0–29.9 | 137 (22.7) | 3,344 (24.0) | -3.1 | 137 (22.7) | 132 (21.9) | 2.0 |
| ≥30.0 | 46 (7.6) | 1,062 (7.6) | 0.0 | 46 (7.6) | 39 (6.5) | 4.5 |
| Missing data | 63 (10.4) | 2,060 (14.8) | -13.1 | 63 (10.4) | 59 (9.8) | 2.2 |
| Japan Coma Scale upon admission, *n* (%) |  |  |  |  |  |  |
| 0 (alert) | 310 (51.4) | 6,153 (44.2) | 14.5 | 310 (51.4) | 283 (46.9) | 9.0 |
| 1–3 (dizzy) | 84 (13.9) | 1,159 (8.3) | 17.9 | 84 (13.9) | 91 (15.1) | -3.3 |
| 10–30 (somnolent) | 27 (4.5) | 560 (4.0) | 2.3 | 27 (4.5) | 25 (4.1) | 1.6 |
| 100–300 (coma) | 182 (30.2) | 6,050 (43.5) | -27.8 | 182 (30.2) | 204 (33.8) | -7.8 |
| Charlson comorbidity index score, mean (SD) | 1.1 (1.2) | 1.2 (1.3) | -2.4 | 1.1 (1.2) | 1.3 (1.3) | -8.8 |
| Comorbidity of peripheral vascular diseases, *n* (%) | 16 (2.7) | 438 (3.1) | -2.9 | 16 (2.7) | 16 (2.7) | 0 |
| Physical function upon admission, *n* (%) |  |  |  |  |  |  |
| Total/severe dependence (BI 0-60) | 409 (67.8) | 8,630 (62.0) | 12.3 | 409 (67.8) | 402 (66.7) | 2.5 |
| Slight/moderate dependence (BI 61-99) | 13 (2.2) | 385 (2.8) | -3.9 | 13 (2.2) | 16 (2.7) | -3.2 |
| Independent (BI = 100) | 86 (14.3) | 2,617 (18.8) | -12.2 | 86 (14.3) | 77 (12.8) | 4.4 |
| Missing | 95 (15.8) | 2,290 (16.4) | -1.9 | 95 (15.8) | 108 (17.9) | -5.8 |
| Cognitive function before admission, *n* (%) |  |  |  |  |  |  |
| No dementia | 403 (66.8) | 10,446 (75.0) | -18.1 | 403 (66.8) | 398 (66.0) | 1.8 |
| Mild dementia | 31 (5.1) | 770 (5.5) | -1.7 | 31 (5.1) | 33 (5.5) | -1.5 |
| Moderate/severe dementia | 21 (3.5) | 469 (3.4) | 0.6 | 21 (3.5) | 29 (4.8) | -6.7 |
| Missing | 148 (24.5) | 2,237 (16.1) | 21.2 | 148 (24.5) | 143 (23.7) | 1.9 |
| Home medical care before admission, *n* (%) | 5 (0.8) | 172 (1.2) | -4.0 | 5 (0.8) | 10 (1.7) | -7.5 |
| Place before admission, *n* (%) |  |  |  |  |  |  |
| Home | 473 (78.4) | 12,736 (91.5) | -37.1 | 473 (78.4) | 493 (81.8) | -8.3 |
| Other hospitals | 124 (20.6) | 1,081 (7.8) | 37.3 | 124 (20.6) | 99 (16.4) | 10.7 |
| Nursing home | 6 (1.0) | 105 (0.8) | 2.6 | 6 (1.0) | 11 (1.8) | -7.0 |
| Ambulance use, *n* (%) | 474 (78.6) | 10,512 (75.5) | 7.4 | 474 (78.6) | 487 (80.8) | -5.4 |
| Primary diagnosis, *n* (%) |  |  |  |  |  |  |
| Acute coronary syndrome | 335 (55.6) | 7,846 (56.4) | -1.6 | 335 (55.6) | 341 (56.6) | -2.0 |
| Cardiac arrest | 94 (15.6) | 3,570 (25.6) | -25.0 | 94 (15.6) | 115 (19.1) | -9.2 |
| Ventricular tachycardia or fibrillation | 50 (8.3) | 1,586 (11.4) | -10.4 | 50 (8.3) | 49 (8.1) | 0.6 |
| Heart failure | 85 (14.1) | 1,545 (11.1) | 9.0 | 85 (14.1) | 93 (15.4) | -3.7 |
| Valve disease | 27 (4.5) | 761 (5.5) | -4.5 | 27 (4.5) | 22 (3.6) | 4.2 |
| Myocarditis | 74 (12.3) | 629 (4.5) | 28.2 | 74 (12.3) | 74 (12.3) | 0 |
| Cardiomyopathy | 46 (7.6) | 495 (3.6) | 17.8 | 46 (7.6) | 44 (7.3) | 1.3 |
| Aortic disease | 13 (2.2) | 193 (1.4) | 5.8 | 13 (2.2) | 10 (1.7) | 3.6 |
| Extracorporeal cardiopulmonary resuscitation, *n* (%) | 139 (23.1) | 6,221 (44.7) | -46.9 | 139 (23.1) | 140 (23.2) | -0.4 |
| Interventions before ECMO, *n* (%) |  |  |  |  |  |  |
| Percutaneous coronary intervention | 306 (50.7) | 7,616 (54.7) | -7.9 | 306 (50.7) | 313 (51.9) | -2.3 |
| Coronary artery bypass grafting | 34 (5.6) | 804 (5.8) | -0.6 | 34 (5.6) | 39 (6.5) | -3.5 |
| Surgical valve procedures | 22 (3.6) | 439 (3.2) | 2.7 | 22 (3.6) | 17 (2.8) | 4.7 |
| Percutaneous valve procedures | 4 (0.7) | 219 (1.6) | -8.7 | 4 (0.7) | 3 (0.5) | 2.2 |
| Organ failure supports on ECMO initiation, *n* (%) |  |  |  |  |  |  |
| Red blood cell transfusion | 449 (74.5) | 8,647 (62.1) | 26.8 | 449 (74.5) | 443 (73.5) | 2.3 |
| Fresh frozen plasma transfusion | 349 (57.9) | 6,634 (47.7) | 20.6 | 349 (57.9) | 354 (58.7) | -1.7 |
| Platelet transfusion | 147 (24.4) | 2,175 (15.6) | 22.0 | 147 (24.4) | 134 (22.2) | 5.1 |
| Dopamine | 123 (20.4) | 4,701 (33.8) | -30.4 | 123 (20.4) | 117 (19.4) | 2.5 |
| Dobutamine | 313 (51.9) | 6,191 (44.5) | 14.9 | 313 (51.9) | 299 (49.6) | 4.6 |
| Noradrenaline | 467 (77.4) | 10,654 (76.5) | 2.2 | 467 (77.4) | 462 (76.6) | 2.0 |
| Adrenaline | 314 (52.1) | 9,802 (70.4) | -38.3 | 314 (52.1) | 308 (51.1) | 2.0 |
| Vasopressin | 39 (6.5) | 982 (7.1) | -2.3 | 39 (6.5) | 43 (7.1) | -2.6 |
| Renal replacement therapy | 124 (20.6) | 2,596 (18.6) | 4.8 | 124 (20.6) | 121 (20.1) | 1.2 |
| Hospital characteristics |  |  |  |  |  |  |
| Teaching hospital, *n* (%) | 594 (98.5) | 13,676 (98.2) | 2.2 | 594 (98.5) | 593 (98.3) | 1.3 |
| Tertiary emergency hospital, *n* (%) | 443 (73.5) | 8,802 (63.2) | 22.2 | 443 (73.5) | 429 (71.1) | 5.2 |
| Annual hospital volume of ECMO, mean (SD) | 69.4 (28.1) | 52.5 (30.9) | 57.2 | 69.4 (28.1) | 70.1 (33.3) | -2.4 |

BI, Barthel Index; ECMO, extracorporeal membrane oxygenation; ECPella, combination of ECMO and Impella; IABP, intra-aortic balloon pump; SD, standard deviation; SMD, standardized mean difference.

All analyses were performed using STATA/SE 17.0 software (StataCorp). Continuous variables are presented as the mean and standard deviation, and categorical variables are presented as the number and percentage. We considered all reported p-values as two-sided and a p<0.05 as statistically significant.

### Sensitivity analysis

We performed two sensitivity analyses. First, we performed an overlap weighting analysis.^23^ The overlap weighting analysis emphasized the target population with the most overlap in the observed characteristics between two groups. The differences and their 95% confidence intervals were calculated with weighted generalized linear models to compare the outcomes. Second, patients who received ECPR may have had a substantially different clinical course in terms of post-cardiac arrest syndrome and the prognosis as compared to those with cardiogenic shock who did not receive ECPR. Therefore, we performed sensitivity analyses excluding the patients who received ECPR.

## Results

### Patient characteristics and outcomes

During the study period, we identified 14,525 eligible patients from 661 hospitals (**Figure 1**). Of those, 603 (4.2%) were identified as the ECPella group, and 13,922 (96%) as the ECMO+IABP group. Of the overall 661 hospitals, Impella was used in 155 (23%).

**Figure 1.**
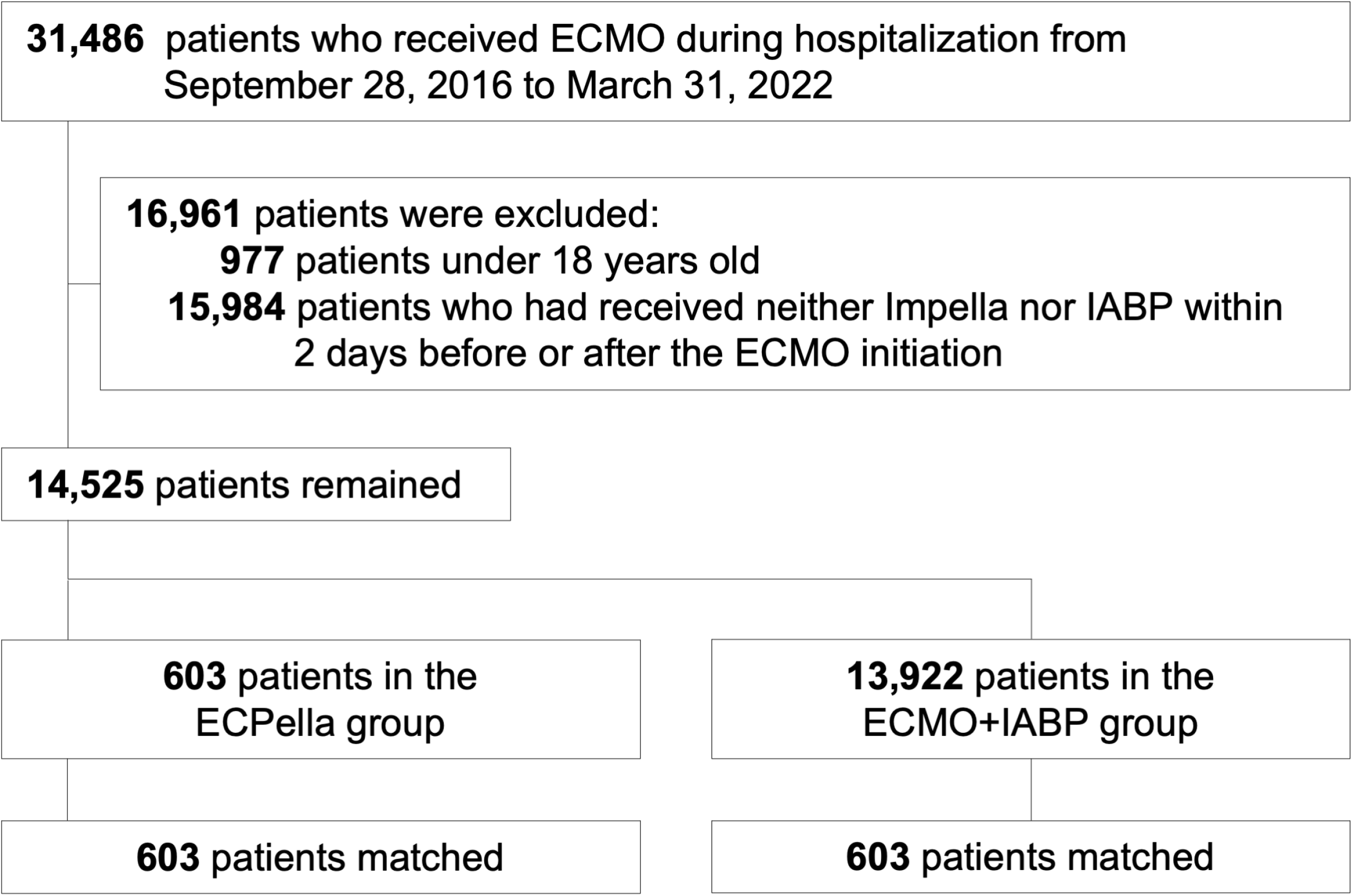
Patient flowchart ECMO, extracorporeal membrane oxygenation; ECPella, combination of ECMO and Impella; IABP, intra-aortic balloon pump.

**Table 1** shows the patient characteristics before and after propensity score matching. In the overall cohort, the mean age was 65 years, and 76% were men. Before the propensity score matching, the patients in the ECPella group tended to be younger, had a poor physical and cognitive function upon admission, were transferred from another hospital, had cardiomyopathy, received blood transfusions and dobutamine, and were admitted to a tertiary emergency hospital with a high annual hospital volume of ECMO. In contrast, the patients in the ECMO+IABP group tended to have poor consciousness, were admitted from home, had a cardiac arrest and ventricular tachycardia or fibrillation, and required ECPR, dopamine, and adrenaline. One-to-one propensity score matching created 603 matched pairs. The distributions of the propensity scores before and after the matching are shown in **Supplementary Figures S1** and **S2**. After the propensity score matching, the patient characteristics were well-balanced between the two groups (**Table 1**).

**Table 2** shows the outcomes before and after propensity score matching. The crude in-hospital mortality was 58.9% in the ECPella group and 65.6% in the ECMO+IABP group. After propensity score matching, there was no significant difference in in-hospital mortality between the ECPella and ECMO+IABP groups (58.9% versus 56.6%; risk difference, 2.3%; 95% confidence interval, −3.9% to 8.5%). A Kaplan–Meier analysis with the log-rank test showed no significant difference in the 60-day in-hospital mortality between the two groups (P value = 0.114) (**Figure 2**). The length of hospital stay was significantly longer in the ECPella group (mean 42.8 days versus 33.7 days; risk difference, 9.1 days; 95% confidence interval, 2.6 to 15.6 days). The total hospitalization cost was also significantly higher in the ECPella group than in the ECMO+IABP group (12,573,000 yen versus 6,857,000 yen; risk differences, 5,716,000 yen; 95% confidence interval, 4,439,000 yen to 6,993,000 yen). Complications more frequently occurred in the ECPella group than in the ECMO+IABP group, including major bleeding (4.0% versus 2.0%; risk difference, 2.0%; 95% confidence interval, 0.007% to 4.0%), vascular complications (4.1% versus 2.2%; risk difference, 2.0%; 95% confidence interval, 0.04% to 3.9%), and renal replacement therapy during hospitalization (50.6% versus 41.1%; risk difference, 9.5%; 95% confidence interval, 3.0% to 15.9%). There were no significant differences in the other secondary outcomes including the length of ECMO (mean 3.9 days versus 3.3 days; risk difference, 0.6 days; 95% confidence interval, −0.8 to 2.0 days), and ischemic strokes (4.1% versus 4.1%; risk difference, 0.0%; 95% confidence interval, −2.1% to 2.1%). The results of the sensitivity analyses, using an overlap weighting analysis, showed similar results to those for the main analyses (**Table 3**).

**Figure 2.**
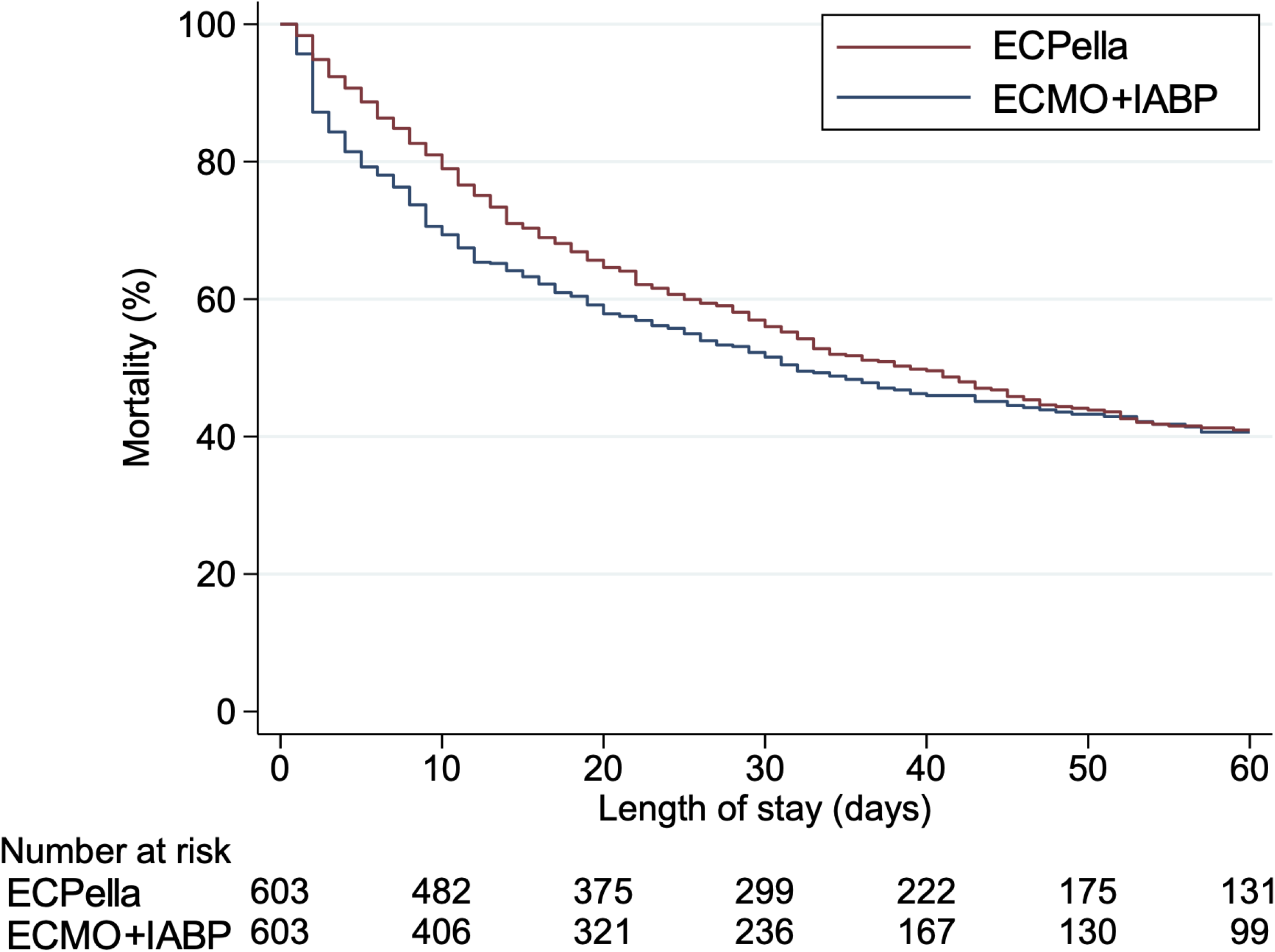
Kaplan–Meier curves for the 60-day in-hospital mortality ECMO, extracorporeal membrane oxygenation; ECPella, combination of ECMO and Impella; IABP, intra-aortic balloon pump.

**Table 2.**
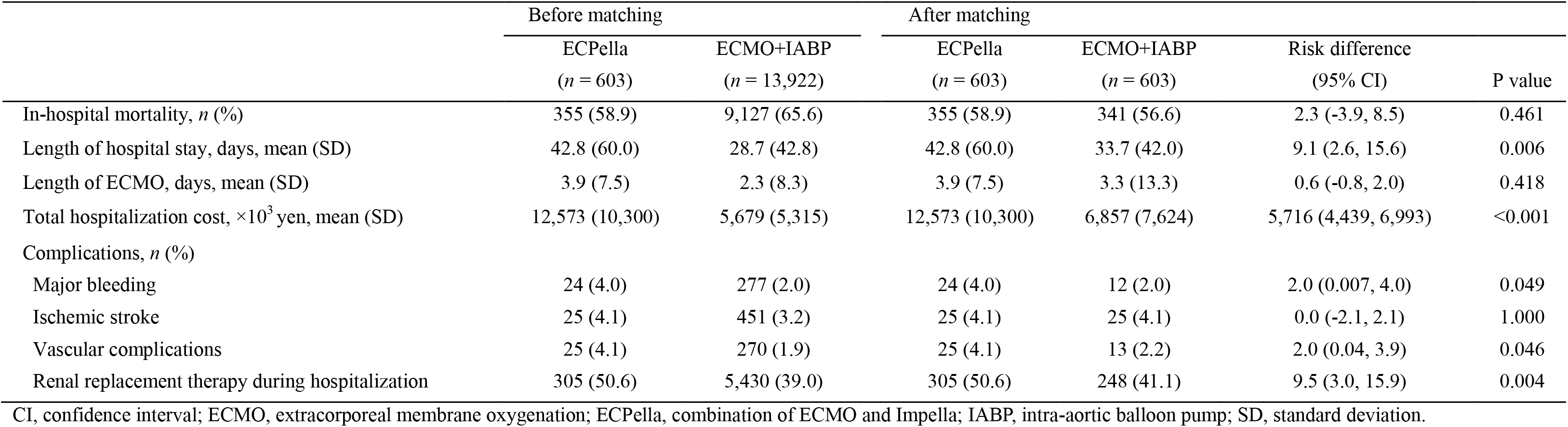
Outcomes before and after propensity score matching.

|  | Before matching |  | After matching |  | Risk difference<br>(95% CI) | P value |
| --- | --- | --- | --- | --- | --- | --- |
|  | ECPella<br>(*n* = 603) | ECMO+IABP<br>(*n* = 13,922) | ECPella<br>(*n* = 603) | ECMO+IABP<br>(*n* = 603) |  |  |
| In-hospital mortality, *n* (%) | 355 (58.9) | 9,127 (65.6) | 355 (58.9) | 341 (56.6) | 2.3 (-3.9, 8.5) | 0.461 |
| Length of hospital stay, days, mean (SD) | 42.8 (60.0) | 28.7 (42.8) | 42.8 (60.0) | 33.7 (42.0) | 9.1 (2.6, 15.6) | 0.006 |
| Length of ECMO, days, mean (SD) | 3.9 (7.5) | 2.3 (8.3) | 3.9 (7.5) | 3.3 (13.3) | 0.6 (-0.8, 2.0) | 0.418 |
| Total hospitalization cost, ×10<sup>3</sup> yen, mean (SD) | 12,573 (10,300) | 5,679 (5,315) | 12,573 (10,300) | 6,857 (7,624) | 5,716 (4,439, 6,993) | <0.001 |
| Complications, *n* (%) |  |  |  |  |  |  |
| Major bleeding | 24 (4.0) | 277 (2.0) | 24 (4.0) | 12 (2.0) | 2.0 (0.007, 4.0) | 0.049 |
| Ischemic stroke | 25 (4.1) | 451 (3.2) | 25 (4.1) | 25 (4.1) | 0.0 (-2.1, 2.1) | 1.000 |
| Vascular complications | 25 (4.1) | 270 (1.9) | 25 (4.1) | 13 (2.2) | 2.0 (0.04, 3.9) | 0.046 |
| Renal replacement therapy during hospitalization | 305 (50.6) | 5,430 (39.0) | 305 (50.6) | 248 (41.1) | 9.5 (3.0, 15.9) | 0.004 |

CI, confidence interval; ECMO, extracorporeal membrane oxygenation; ECPella, combination of ECMO and Impella; IABP, intra-aortic balloon pump; SD, standard deviation.

**Table 3.** Overlap weighting.

|  | After IPTW |  |  |  |
| --- | --- | --- | --- | --- |
|  | ECPella | ECMO+IABP | Risk difference (95% CI) | P value |
| In-hospital mortality, % | 60.1 | 57.8 | 2.4 (-2.3, 7.0) | 0.324 |
| Length of hospital stay, days, mean (SD) | 41.1 (56.7) | 33.9 (48.8) | 7.2 (2.1, 12.4) | 0.006 |
| Length of ECMO, days, mean (SD) | 3.8 (7.2) | 2.9 (12.2) | 0.9 (0.06, 1.8) | 0.035 |
| Total hospitalization cost, $\times 10^3$ yen, mean (SD) | 12,133 (9,472) | 6,758 (7,014) | 5,375 (4,310, 6,440) | <0.001 |
| Complications, % |  |  |  |  |
| Major bleeding | 3.9 | 2.5 | 1.5 (-0.2, 3.2) | 0.091 |
| Ischemic stroke | 4.1 | 3.8 | 0.3 (-1.3, 1.9) | 0.709 |
| Vascular complications | 4.1 | 2.4 | 1.7 (-0.2, 3.5) | 0.078 |
| Renal replacement therapy during hospitalization | 49.8 | 40.5 | 9.3 (4.2, 14.5) | <0.001 |

CI, confidence interval; ECMO, extracorporeal membrane oxygenation; ECPella, combination of ECMO and Impella; IABP, intra-aortic balloon pump; SD, standard deviation.

**Supplementary Table S2.** shows the results of the sensitivity analyses excluding patients who received ECPR. After excluding 6,360 patients (44%) who received ECPR, there was no significant difference in in-hospital mortality between the ECPella and ECMO+IABP groups (57.3% versus 50.9%; risk difference, 6.5%; 95% confidence interval, −0.2% to 13.2%). The results of the secondary outcomes were also consistent with the main analyses.

## Discussion

To the best of our knowledge, this nationwide cohort study had the largest number of patients among the studies comparing ECPella versus ECMO+IABP. There was no significant difference in in-hospital mortality between the ECPella and ECMO+IABP groups. Meanwhile, ECPella was significantly associated with a longer hospital stay, higher total hospitalization cost, and more complications than ECMO+IABP.

The present study found no survival benefit of ECPella as compared to ECMO+IABP. This was consistent with the results of the previous studies.^8,11^ One of those studies, from the Extracorporeal Life Support Organization registry, showed a considerably higher proportion of Impella use in combination with ECMO (33% versus 4.2%).^8^ The previous study performed a propensity score-matching analysis and concluded ECPella as compared to ECMO+IABP was associated with a similar survival in the 560 matched pairs.^8^ Randomized controlled trials comparing the LV unloading strategies are certainly needed to reach any conclusions. To investigate the comparative effectiveness of Impella or IABP use in combination with ECMO, future studies may need to consider the following. First, the etiology of cardiogenic shock may be a key issue. Underlying diseases with severe impairment of the LV contractility, such as fulminant myocarditis, could receive a pathophysiological benefit from ECPella as a bridge to recovery;^24,25^ however, the current number of cases could not substantiate this subgroup analysis. Indeed, the present study included only 140/1,206 (11.6%) patients with myocarditis. Second, the relationship between the procedural volume (i.e., learning curves) and outcomes should also be considered. Data are scarce for Impella, but the association between the procedural volume and outcomes has been shown for ECMO.^26^ Hence, when involving data early after the approval of the Impella, as in the present study, the survival benefit of ECPella may be underestimated. Finally, early recognition of cardiogenic shock and its stabilization by an appropriate timing of mechanical circulatory support (MCS) device use may be more critical than that in which an MCS device is used.^27,28^

In the present study, the total hospitalization cost was approximately 1.8 times higher in the ECPella group than in the ECMO+IABP group. This was partly because the cost of the Impella itself was considerably higher than that of an IABP or ECMO; it was approximately 17 times higher than an IABP and 7 times higher than ECMO, according to the national health reimbursement data in Japan.^29^ Moreover, a longer hospital stay and more complications including major bleeding, vascular complications, and renal replacement therapy during hospitalization may have contributed to a higher total hospitalization cost. A recent study, using the nationwide inpatient database in Japan, that included patients who required MCS early after admission also showed higher medical costs for ECPella than ECMO+IABP, potentially due to more frequent blood transfusions, a longer duration of ventilator support, and a longer length of hospital stay.^30^ In previous studies focusing on the temporal trends before and after the approval of the Impella, the hospitalization cost in the Impella era was higher than that in the pre-Impella era, especially in hospitals where the Impella was more frequently used.^31,32^ Unplanned readmissions may be an important quality indicator in patients who received an Impella, with limited previous studies showing a high incidence of a 30-day readmission of more than 10%.^33^ Unless the survival benefit and reduction in readmissions for ECPella as compared to ECMO+IABP is confirmed, clinicians should be cautious about the patient selection for Impella use in combination with ECMO for LV unloading.

Our results included some important clinical implications. First, the present study provided no evidence of whether IABP or Impella was better for use in combination with ECMO, and evidence of a longer hospital stay, higher total hospitalization cost, and more complications for ECPella. This suggested that ECPella should be implemented in carefully selected patients. Second, further studies are warranted to investigate the outcomes according to the etiology of the cardiogenic shock, learning curves for Impella use, and readmissions in patients who received the Impella.

The present study had several limitations. First, the decision on whether to use an IABP or Impella was at the individual clinician’s discretion due to the nature of the present study using the observational database, which led to confounding by indication. We attempted to control for the measured confounding factors using the propensity score analyses; however, we were unable to control for any possible unmeasured variables, such as the vital signs, laboratory data, or LV function. Second, the present study was unable to identify whether patients initially received IABP or Impella and then ECMO, or whether they initially received ECMO and then IABP or Impella for LV unloading. In addition, we defined 152 patients (40%) who received Impella and IABP in combination with ECMO on the same day as the ECPella group; however, some patients might have been downgraded from Impella to IABP due to Impella-related complications. Therefore, misclassifications may have led to a bias in our study. The order and combination of the MCS devices depended on the changing severity of the cardiogenic shock, suggesting that it would be difficult to accurately categorize those complex processes even if additional information were available during hospitalization. Third, the present study was also unable to identify whether the patients received an Impella 2.5, Impella CP, or Impella 5.0 in the ECPella group. Fourth, the incidence of complications in the present study was considerably lower than in the previous studies.^8,11^ Given that the sensitivity of the diagnosis might have been low in our database, there was a possibility of underreporting complications. Finally, many patients in the present study received vasoactive drugs including dopamine and adrenaline. That might have delayed the MCS initiation and caution should be taken in interpreting our results.

## Conclusions

The present study using a nationwide inpatient administrative database showed that ECPella was not associated with any survival benefit but was associated with a longer hospital stay, higher total hospitalization cost, and more complications as compared to ECMO+IABP.

## Declarations

### Ethics approval and consent to participate

This study was performed in accordance with the amended Declaration of Helsinki, and the Institutional Review Board of The University of Tokyo approved this study (approval number: 3501-(3); 25 December 2017).

### Consent for publication

The review board waived the requirement for informed consent because of the anonymous nature of the data. No information describing the individual patients, hospitals, or treating physicians was obtained.

### Availability of data and materials

The datasets analyzed during the current study are not publicly available owing to contracts with the hospitals providing the data to the database.

### Authors’ contributions

**Yuji Nishimoto**: Conceptualization, Software, Formal analysis, Investigation, Writing - Original Draft; **Hiroyuki Ohbe**: Conceptualization, Methodology, Software, Formal analysis, Investigation, Writing - Review and Editing; **Jun Nakata**: Conceptualization, Methodology, Investigation, Writing - Review and Editing, Supervision; **Toru Takiguchi**: Conceptualization, Methodology, Investigation, Writing - Review and Editing, Supervision; **Mikio Nakajima**: Conceptualization, Investigation, Writing - Review and Editing, Supervision; **Yusuke Sasabuchi**: Conceptualization, Investigation, Writing - Review and Editing, Supervision; **Toshiaki Isogai**: Software, Formal analysis, Investigation, Data Curation; **Hiroki Matsui**: Software, Formal analysis, Investigation, Data Curation; **Yukihito Sato:** Software, Investigation, Resources, Supervision; **Tetsuya Watanabe**: Software, Investigation, Resources, Supervision; **Takahisa Yamada**: Software, Investigation, Resources, Supervision; **Masatake Fukunami**: Software, Investigation, Resources, Supervision; **Hideo Yasunaga**: Writing - Review and Editing, Supervision. All authors read and approved the final manuscript.

## Acknowledgments

We would like to express our gratitude to Mr. John Martin for his grammatical assistance.

